# Subjective Financial Strain and Incident Heart Disease Among US Adults Aged 50 Years or Older

**DOI:** 10.64898/2026.02.23.26346937

**Authors:** Derek Tharp

## Abstract

**Background:** Financial strain has been linked to adverse cardiovascular outcomes, yet whether this association persists beyond objective socioeconomic resources remains unclear. We examined associations of financial strain with incident heart disease and all-cause mortality among US adults aged 50 years or older.

**Methods:** Prospective cohort study using the Health and Retirement Study (2006-2022). Among 7219 participants completing the Psychosocial Leave-Behind Questionnaire, the exposure was ongoing financial strain (high vs low/none). Incident heart disease was assessed among 4956 participants without baseline cardiovascular disease using cause-specific Cox and Fine-Gray models. All-cause mortality was modeled using sequential Cox regression.

**Results:** Among 7219 participants (mean [SD] age, 67.5 [10.6] years; 58.6% female), 1423 (19.7%) reported high financial strain. Financial strain was associated with incident heart disease (cause-specific HR, 1.18; 95% CI, 1.02-1.37; *P* =.03; 1310 events), corroborated by Fine-Gray models (SHR, 1.16; 95% CI, 1.00-1.34). For all-cause mortality (3466 deaths), financial strain was associated after demographic and clinical adjustment (HR, 1.17; 95% CI, 1.07-1.28) but attenuated after further adjustment for income and wealth (HR, 1.10; 95% CI, 1.00-1.20; *P* =.051). The mortality association differed by age (interaction *P* =.001): HR, 1.25 (95% CI, 1.03-1.52) for adults younger than 65 years versus HR, 1.04 (95% CI, 0.94-1.16) for those 65 or older.

**Conclusions:** Financial strain was associated with incident heart disease independent of socioeconomic resources. The mortality association was attenuated by income and wealth adjustment but remained elevated among preretirement adults. Financial strain may be a clinically accessible marker of cardiovascular risk among working-age adults.

## CLINICAL PERSPECTIVE

### What Is New?

- In a prospective cohort of 4956 adults without baseline cardiovascular disease followed for up to 16 years, subjective financial strain was associated with 18% higher risk of incident heart disease (cause-specific HR, 1.18; 95% CI, 1.02-1.37) after adjustment for demographics, health behaviors, clinical conditions, income, and wealth.
- The financial strain-mortality association was significantly modified by age (P for interaction =.001), with a 25% higher hazard among adults younger than 65 years (HR, 1.25; 95% CI, 1.03-1.52) but no significant association among those 65 years or older.
- The association of financial strain with all-cause mortality was attenuated after adjustment for objective socioeconomic resources (41% reduction in excess risk), whereas the heart disease association persisted after the same adjustment.

### What Are the Clinical Implications?

- A single screening question about ongoing financial strain could help identify patients at elevated cardiovascular risk, particularly among working-age adults not yet eligible for Medicare.
- Clinicians conducting cardiovascular risk assessment may consider financial strain as a marker that prompts evaluation of cost-related medication nonadherence, forgone preventive care, and food insecurity.
- The mortality association was concentrated among adults younger than 65 years (HR, 1.25; 95% CI, 1.03-1.52), a population facing greater income volatility and less comprehensive health coverage, supporting targeted screening during annual wellness visits for preretirement adults.

## Introduction

Financial strain—the subjective experience of difficulty meeting financial obligations—is a psychosocial factor associated with cardiovascular morbidity and mortality.^1,2^ Unlike objective measures of socioeconomic status such as income and wealth, financial strain captures the individual’s appraisal of resource inadequacy, a dimension of economic stress that standard socioeconomic indicators do not directly measure.^3^ Whether this subjective appraisal is associated with cardiovascular outcomes after adjustment for objective socioeconomic position, or primarily reflects confounding by material resources, has implications for cardiovascular risk assessment.

Chronic financial strain may contribute to allostatic load through sustained hypothalamic-pituitary-adrenal axis activation and inflammatory signaling,^4,5,6^ though whether subjective appraisal of financial difficulty activates these pathways beyond the material deprivation it indexes is unknown. Financial strain may also operate through behavioral intermediates (eg, forgoing care or medications because of cost), although these were not directly assessed in the present study.^7^

Prior investigations using the Health and Retirement Study (HRS) have linked financial measures to cardiovascular and mortality outcomes. Pool et al^8^ found that a 75% or greater loss in net worth was associated with a 50% increase in mortality risk. Tucker-Seeley et al^9^ documented associations between material hardship and mortality. Machado et al^10^ demonstrated that midlife wealth mobility was associated with long-term cardiovascular health, with downward wealth trajectories linked to worse cardiovascular outcomes. However, these studies used objective financial measures and did not examine whether subjective financial strain is associated with cardiovascular outcomes after adjustment for income and wealth.

We addressed this question using 16 years of follow-up data from the HRS, examining the association of subjective financial strain with incident heart disease and all-cause mortality. We tested whether these associations persisted after adjustment for objective socioeconomic resources, whether they differed by age group, and whether a dose-response gradient was present.

## Methods

### Study Design and Population

We conducted a prospective cohort study using data from the Health and Retirement Study (HRS), a nationally representative longitudinal survey of US adults aged 50 years and older and their spouses.^11^ The HRS began in 1992 and conducts biennial interviews. We used the RAND HRS Longitudinal File (2022, Version 1) for harmonized variables and the HRS cross-wave tracker file for mortality ascertainment. Since 2006, the Psychosocial Leave-Behind Questionnaire (LBQ) has been administered to an alternating random 50% subsample of respondents who complete the enhanced face-to-face interview (EFTF) each wave, such that individuals complete the LBQ approximately every 4 years; the LBQ includes measures of chronic stressors. Eligibility for the LBQ is therefore conditional on EFTF completion, which may introduce selection if EFTF participation differs by financial strain or health status.

Our baseline cohort included participants who (1) completed the 2006 wave (Wave 8) interview, (2) were assigned and returned the LBQ with valid responses to the financial strain item, and (3) were aged 50 years or older at baseline (*N* = 7219). For regression analyses, we used complete-case analysis for covariates included in each model; the primary mortality model included 7064 participants with complete covariate data (Table 3). For incident heart disease analyses, we further restricted to 4956 participants without self-reported heart disease or stroke at baseline and with complete covariate data. Follow-up continued through 2022 (Wave 16).

Participant flow is shown in Figure S1; comparison of included versus excluded participants and LBQ responders versus nonresponders is shown in Tables S7 and S9.

The HRS protocol is approved by the University of Michigan Institutional Review Board, and all participants provided written informed consent. This secondary analysis used publicly available, deidentified data and was deemed exempt from review.

### Exposure: Subjective Financial Strain

Financial strain was assessed using the “Ongoing financial strain” item from the HRS Psychosocial Leave-Behind Questionnaire (LBQ) Ongoing Chronic Stressors module, which asks respondents to report current and ongoing problems that have lasted 12 months or longer and, if present, how upsetting the problem has been: (1) No, did not happen; (2) Yes, but not upsetting; (3) Yes, somewhat upsetting; (4) Yes, very upsetting.^12^ This item captures both exposure to an ongoing financial stressor and its appraisal; other HRS financial strain/hardship measures include end-of-month financial strain and material hardship items.

For primary analyses, we created a binary exposure (upsetting financial strain: somewhat/very upsetting vs no/not upsetting). We also examined the 4-level ordinal responses to assess dose-response relationships. We note that the standard HRS chronic stressor index is typically operationalized as the count of stressors rated “yes, very upsetting”; our exposure focused on the financial strain item rather than the multi-item index.

### Outcomes

#### Incident Heart Disease

Among participants without heart disease or stroke at baseline, incident self-reported heart disease (composite outcome) was defined as the first affirmative response to whether a doctor had told the respondent they had “a heart attack, coronary heart disease, angina, congestive heart failure, or other heart problems.” Event time was the interview date of first report (interval-censored by biennial ascertainment). Because death precludes ascertainment, we report both cause-specific Cox models and Fine-Gray subdistribution hazard models.

#### All-Cause Mortality

Mortality status and month/year of death were obtained from the HRS cross-wave tracker file, which integrates National Death Index linkage and proxy/exit interview reports. Follow-up time was calculated from the 2006 interview month/year to death month/year or December 31, 2022, whichever came first.

### Covariates

Covariates were selected a priori based on theoretical associations with financial strain and health outcomes. Demographics included age, sex, race and ethnicity (non-Hispanic White, non-Hispanic Black, Hispanic, non-Hispanic other), and marital status (married or partnered vs other; separated individuals were classified as not partnered). Race and ethnicity were derived from the HRS race and Hispanic ethnicity items; Hispanic ethnicity was prioritized over race when constructing mutually exclusive categories. Socioeconomic factors included years of education and baseline total household income (respondent plus spouse/partner; RAND HRS H8ITOT) and total household net wealth (RAND HRS H8ATOTB), which sums financial and nonfinancial assets (including primary residence equity and IRA/Keogh accounts) and subtracts mortgages and other debts; income and wealth were transformed using the inverse hyperbolic sine to handle zero and negative values. Health behaviors included current smoking status and body mass index (BMI; calculated as weight in kilograms divided by height in meters squared). Baseline health conditions included self-reported physician-diagnosed hypertension, diabetes, heart disease, and stroke.

Race and ethnicity were included because of known disparities in both financial strain and health outcomes; these data were self-reported by participants using HRS standard categories.

Because exposure and covariates were measured in the same wave, models adjusting for health behaviors, conditions, or depressive symptoms may represent overadjustment for variables on the causal pathway; sequential models should be interpreted as sensitivity analyses to alternative adjustment assumptions. In sensitivity analyses, we additionally adjusted for depressive symptoms (8-item Center for Epidemiologic Studies Depression Scale [CES-D] score ≥3).

## Statistical Analysis

Baseline characteristics were compared by financial strain status using standardized mean differences (SMDs), with absolute SMDs greater than 0.10 indicating meaningful imbalance.^13^

The target estimand is the conditional hazard ratio for financial strain under the assumption of no unmeasured confounding (exchangeability), consistency of the exposure, and positivity across covariate strata. Because this is an observational study, we interpret results as adjusted associations rather than causal effects.

Hazard ratios (HRs) and 95% CIs were estimated using Cox proportional hazards regression with time since the 2006 interview as the time scale and the Efron method for ties. We assessed the proportional hazards assumption using Schoenfeld residuals (global and covariate-specific tests); diagnostics are reported in Table S10. We constructed sequential models recognizing that income and wealth may function as confounders, mediators, or both: Model 1 (unadjusted); Model 2 (age and sex); Model 3 (demographics, behaviors, health conditions); and Model 3b (additionally adjusted for income and wealth; designated as primary). Models 3 and 3b bound the plausible range under different causal assumptions. Model 4 additionally included depressive symptoms; Model 4b additionally included both depressive symptoms and self-rated health.

We performed propensity score matching (1:1 nearest-neighbor, without replacement, caliper of 0.2 SD of logit propensity score) using covariates from the primary model plus baseline self-rated health.^14^ Balance was assessed using SMDs (Table S1; Figure S2). HRs were estimated using Cox regression stratified by matched pair, targeting the average treatment effect on the treated (ATT). E-values were calculated to assess sensitivity to unmeasured confounding.^15^

Effect modification by age was prespecified (<65 vs ≥65 years, approximating Medicare eligibility) using interaction terms and age-stratified models with within-stratum age adjustment. Continuous age interaction was tested using age centered at 65 years.

We calculated absolute mortality risks at 5, 10, and 15 years using Kaplan-Meier methods (Table S8); covariate-standardized absolute risks within age strata were estimated using marginal standardization from fitted Cox models (Table S12). Sensitivity analyses excluded deaths in the first 1 or 2 years to address potential reverse causation. Because the HRS includes spouses, we conducted sensitivity analyses using robust standard errors clustered at the household level to account for within-couple correlation (Table S13).

Analyses were conducted using Python version 3.13 with the lifelines package (version 0.29); Fine-Gray models were estimated using Stata version 18 (stcrreg command). Two-sided P values less than.05 were considered statistically significant for the prespecified primary analysis (Model 3b association of financial strain with all-cause mortality). The analysis of incident heart disease, age interaction, dose-response, propensity score analyses, and other sensitivity/subgroup analyses were considered secondary and were interpreted with caution in light of multiple comparisons (no formal multiplicity adjustment was applied). Primary analyses were unweighted; survey-weighted sensitivity analyses using the HRS LBQ respondent weight (R8LBWGTR) are reported in Table S11. Because the sandwich estimator does not fully account for HRS complex survey design, we retain the unweighted analysis as primary.

## Results

### Study Population

Of 18,469 HRS Wave 8 respondents, 7432 returned the LBQ with valid financial strain data. Of these, 7219 were aged 50 years or older and formed the baseline cohort (Figure S1).

Complete covariate data were available for 7064 participants, who comprised the analytic cohort for the primary mortality models (Table 3). Among the baseline cohort, 1423 (19.7%) reported high financial strain (Table 1). LBQ responders were similar to other Wave 8 respondents on measured characteristics (all absolute SMDs ≤0.14; Table S9).

**Table 1.**
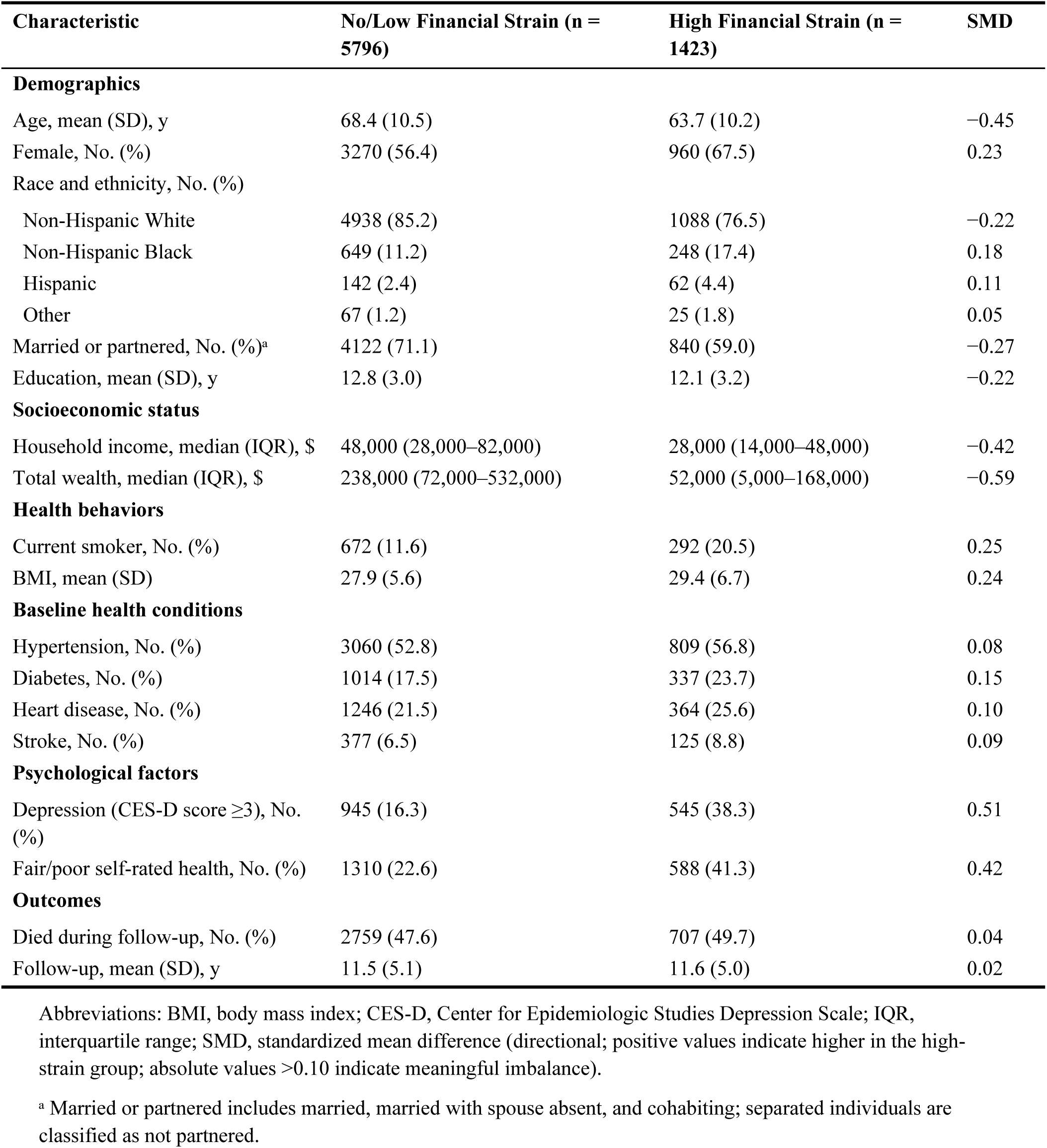
Baseline Characteristics of Study Participants by Financial Strain Status (*N* = 7219)

| Characteristic | No/Low Financial Strain (n = 5796) | High Financial Strain (n = 1423) | SMD |
| --- | --- | --- | --- |
| **Demographics** |  |  |  |
| Age, mean (SD), y | 68.4 (10.5) | 63.7 (10.2) | −0.45 |
| Female, No. (%) | 3270 (56.4) | 960 (67.5) | 0.23 |
| Race and ethnicity, No. (%) |  |  |  |
| Non-Hispanic White | 4938 (85.2) | 1088 (76.5) | −0.22 |
| Non-Hispanic Black | 649 (11.2) | 248 (17.4) | 0.18 |
| Hispanic | 142 (2.4) | 62 (4.4) | 0.11 |
| Other | 67 (1.2) | 25 (1.8) | 0.05 |
| Married or partnered, No. (%)<sup>a</sup> | 4122 (71.1) | 840 (59.0) | −0.27 |
| Education, mean (SD), y | 12.8 (3.0) | 12.1 (3.2) | −0.22 |
| **Socioeconomic status** |  |  |  |
| Household income, median (IQR), \$ | 48,000 (28,000–82,000) | 28,000 (14,000–48,000) | −0.42 |
| Total wealth, median (IQR), \$ | 238,000 (72,000–532,000) | 52,000 (5,000–168,000) | −0.59 |
| **Health behaviors** |  |  |  |
| Current smoker, No. (%) | 672 (11.6) | 292 (20.5) | 0.25 |
| BMI, mean (SD) | 27.9 (5.6) | 29.4 (6.7) | 0.24 |
| **Baseline health conditions** |  |  |  |
| Hypertension, No. (%) | 3060 (52.8) | 809 (56.8) | 0.08 |
| Diabetes, No. (%) | 1014 (17.5) | 337 (23.7) | 0.15 |
| Heart disease, No. (%) | 1246 (21.5) | 364 (25.6) | 0.10 |
| Stroke, No. (%) | 377 (6.5) | 125 (8.8) | 0.09 |
| **Psychological factors** |  |  |  |
| Depression (CES-D score ≥3), No. (%) | 945 (16.3) | 545 (38.3) | 0.51 |
| Fair/poor self-rated health, No. (%) | 1310 (22.6) | 588 (41.3) | 0.42 |
| **Outcomes** |  |  |  |
| Died during follow-up, No. (%) | 2759 (47.6) | 707 (49.7) | 0.04 |
| Follow-up, mean (SD), y | 11.5 (5.1) | 11.6 (5.0) | 0.02 |

Abbreviations: BMI, body mass index; CES-D, Center for Epidemiologic Studies Depression Scale; IQR, interquartile range; SMD, standardized mean difference (directional; positive values indicate higher in the high-strain group; absolute values >0.10 indicate meaningful imbalance).
<sup>a</sup> Married or partnered includes married, married with spouse absent, and cohabiting; separated individuals are classified as not partnered.

**Table 2.** Association of Financial Strain With Incident Heart Disease.

| **Model** | ***N*** | **Events** | **HR (95% CI)** | ***P Value*** |
| --- | --- | --- | --- | --- |
| Cause-specific HR (overall)<sup>a</sup> | 4956 | 1310 | 1.18 (1.02-1.37) | .03 |
| Age <65 years<sup>a</sup> | 2192 | 488 | 1.16 (0.94-1.44) | .16 |
| Age ≥65 years<sup>a</sup> | 2764 | 822 | 1.18 (0.96-1.45) | .12 |

Abbreviations: CI, confidence interval; HR, hazard ratio; SHR, subdistribution hazard ratio.
<sup>a</sup> Cause-specific hazard; death without heart disease treated as censoring ( $n = 1375$ competing deaths). Model adjusted for age, sex, race and ethnicity, education, marital status, smoking, body mass index, hypertension, diabetes, income, and wealth (participants with baseline heart disease and stroke were excluded). Fine-Gray subdistribution hazard (SHR, 1.16; 95% CI, 1.00-1.34; $P = .04$ ) accounting for competing mortality is reported in Table S6.

**Table 3.** Association of Financial Strain With All-Cause Mortality.

| **Model** | **N** | **Deaths** | **HR (95% CI)** | **P Value** |
| --- | --- | --- | --- | --- |
| Model 1: Unadjusted | 7219 | 3466 | 0.95 (0.87-1.03) | .20 |
| Model 2: Age + sex | 7219 | 3466 | 1.40 (1.28-1.52) | <.001 |
| Model 3: + Demographics, behaviors, health conditions<sup>a</sup> | 7064 | 3394 | 1.17 (1.07-1.28) | <.001 |
| Model 3b: + Income + wealth (PRIMARY)<sup>b</sup> | 7064 | 3394 | 1.10 (1.00-1.20) | .051 |
| Model 4: + Depressive symptoms<sup>c</sup> | 6962 | 3331 | 1.06 (0.97-1.16) | .22 |
| Model 4b: + All psychological<sup>d</sup> | 6956 | 3326 | 1.03 (0.94-1.13) | .58 |
| Propensity score-matched<sup>e</sup> | 2632 | 1213 | 1.04 (0.91-1.19) | .56 |
| Excluding deaths in first year | 6940 | 3270 | 1.08 (0.98-1.19) | .10 |
| Excluding deaths in first 2 years | 6697 | 3086 | 1.05 (0.95-1.15) | .37 |

Abbreviations: CI, confidence interval; HR, hazard ratio.
<sup>a</sup> Adjusted for age, sex, race and ethnicity, education, marital status, smoking, body mass index, hypertension, diabetes, heart disease, and stroke (no income/wealth).
<sup>b</sup> Model 3 plus household income and total wealth (inverse hyperbolic sine transformed).
<sup>c</sup> Model 3b plus depressive symptoms (CES-D score $\geq 3$ ).
<sup>d</sup> Model 3b plus depressive symptoms and self-rated health.
<sup>e</sup> 1:1 nearest-neighbor matching with caliper (0.2 SD of logit propensity score); covariates aligned with Model 3b plus baseline self-rated health; 1316 matched pairs.

Participants with high financial strain were younger (mean age, 63.7 vs 68.4 years; SMD, −0.45), more likely to be female (67.5% vs 56.4%; SMD, 0.23), more likely to be non-Hispanic Black (17.4% vs 11.2%; SMD, 0.18), less likely to be married or partnered (59.0% vs 71.1%; SMD, −0.27), and had substantially lower household income (SMD, −0.42) and wealth (SMD, −0.59). They also had higher prevalences of current smoking (20.5% vs 11.6%; SMD, 0.25), depression (38.3% vs 16.3%; SMD, 0.51), and fair or poor self-rated health (41.3% vs 22.6%; SMD, 0.42).

### Incident Heart Disease

Among 4956 participants without self-reported heart disease or stroke at baseline, 1310 (26.4%) developed incident self-reported heart disease during follow-up. An additional 1375 participants died without developing heart disease (competing event).

In cause-specific models (treating death as censoring), financial strain was associated with higher risk of incident heart disease (HR, 1.18; 95% CI, 1.02-1.37; *P* =.03) (Table 2). In Fine-Gray subdistribution hazard models accounting for competing mortality, the association was slightly attenuated but remained significant (subdistribution HR, 1.16; 95% CI, 1.00-1.34; *P* =.04; Table S6). Unlike mortality (see below), no significant age interaction was observed for incident heart disease, which argues against a cardiovascular mechanism as the sole explanation for the mortality age interaction.

### All-Cause Mortality

During a mean (SD) follow-up of 11.5 (5.1) years (83,157 total person-years), 3466 deaths (48.0%) occurred.

Despite substantially worse risk factor profiles, participants with high financial strain had similar crude mortality (49.7% vs 47.6%) owing to their younger age distribution (mean 63.7 vs 68.4 years). In unadjusted analysis, financial strain was not significantly associated with mortality (HR, 0.95; 95% CI, 0.87-1.03; *P* =.20), reflecting this strong confounding by age (Table 3). After adjustment for age and sex, financial strain was associated with higher mortality (HR, 1.40; 95% CI, 1.28-1.52; *P* <.001).

After adjustment for demographics, behaviors, and health conditions (Model 3, excluding income and wealth), financial strain remained significantly associated with mortality (HR, 1.17; 95% CI, 1.07-1.28; *P* <.001). In the primary model (Model 3b, additionally adjusted for income and wealth), the association was attenuated and no longer statistically significant (HR, 1.10; 95% CI, 1.00-1.20; *P* =.051), representing a 41% reduction in excess risk. When additionally adjusting for depressive symptoms (Model 4), the association was further attenuated (HR, 1.06; 95% CI, 0.97-1.16; *P* =.22); adjusting for both depressive symptoms and self-rated health (Model 4b) yielded HR, 1.03 (95% CI, 0.94-1.13; *P* =.58).

Propensity score matching created 1316 matched pairs (1316 of 1423 high-strain participants matched) with excellent covariate balance (all post-matching SMDs <0.10; Table S1). In the matched sample, financial strain was not significantly associated with mortality (HR, 1.04; 95% CI, 0.91-1.19; *P* =.56); the point estimate was similar to the fully adjusted regression estimate (Model 3b HR, 1.10), with wider uncertainty reflecting the reduced effective sample size and the ATT estimand restricted to matched participants.

### Effect Modification by Age

In the primary analytic cohort with complete covariate data (*N* = 7064), the association between financial strain and mortality differed significantly by age (*P* for interaction =.001).

Among participants younger than 65 years (*n* = 2660, 568 deaths), financial strain was associated with 25% higher mortality (HR, 1.25; 95% CI, 1.03-1.52; *P* =.02). Among those 65 years or older (*n* = 4404, 2826 deaths), no significant association was observed (HR, 1.04; 95% CI, 0.94-1.16; *P* =.44) (Figure S3).

Continuous age interaction testing (age centered at 65 years) confirmed this pattern (*P* for interaction <.001). The estimated HR decreased with age: HR, 1.43 (95% CI, 1.21-1.70) at age 55; HR, 1.21 (95% CI, 1.09-1.35) at age 65; and HR, 0.95 (95% CI, 0.83-1.07) at age 80 (Table S5).

In contrast, no significant age interaction was observed for incident heart disease (Table 2), which argues against a cardiovascular mechanism as the sole explanation for the mortality age interaction.

### Dose-Response Analysis

When examining the 4-level financial strain measure, no clear dose-response pattern emerged (Table S2). Compared with no strain, reporting strain that was “somewhat upsetting” was associated with higher mortality (HR, 1.12; 95% CI, 1.01-1.24; *P* =.03), while “not upsetting” strain (HR, 0.99; *P* =.86) and “very upsetting” strain (HR, 1.00; 95% CI, 0.85-1.19; *P* =.96) were not associated with mortality. The nonmonotonic pattern and nonsignificant test for linear trend (*P* =.18) raise questions about construct validity or suggest heterogeneity in the small “very upsetting” subgroup (*n* = 373).

### Absolute Risk

At 5 years, unadjusted mortality was 12.5% among those with low/no financial strain versus 14.0% among those with high strain (absolute risk difference, 1.6 percentage points). At 10 years, corresponding figures were 30.1% versus 29.0% (risk difference, −1.1 percentage points), reflecting confounding by age (Table S8). Unadjusted longer-term absolute risks were strongly confounded by baseline age differences.

### Sensitivity Analyses

The CI-bound E-value for the primary estimate (HR, 1.10) was 1.00, meaning that sampling variability alone is sufficient to shift the confidence interval to include the null; no unmeasured confounding is required. For the under-65 age stratum (HR, 1.25), the E-value was 1.82 (CI-bound E-value, 1.22), suggesting somewhat greater robustness (Table S3). Given plausible unmeasured confounders (eg, childhood socioeconomic conditions, personality/negative affectivity, and financial literacy), residual confounding remains a credible alternative explanation.

Excluding deaths in the first year yielded a similar estimate (HR, 1.08; 95% CI, 0.98-1.19; *P* =.10). Excluding deaths in the first 2 years yielded a slightly attenuated estimate (HR, 1.05; 95% CI, 0.95-1.15; *P* =.37). The attenuation with longer exclusion windows most likely reflects progressive exposure misclassification, as a single-timepoint measure of financial strain becomes increasingly uninformative over a 16-year follow-up that spans the 2008 Great Recession. We cannot exclude contributions from reverse causation (preclinical illness increasing perceived strain) or a genuinely time-limited early effect, but the misclassification explanation is most parsimonious given the measurement structure.

Proportional hazards diagnostics indicated no significant violation for financial strain (Schoenfeld residual *P* =.18), though age (*P* <.001), BMI (*P* <.001), and baseline heart disease (*P* =.002) showed nonproportionality (Table S10). To address the age nonproportionality, we note that our age-stratified models effectively relax this assumption and yield consistent conclusions.

LBQ-weighted analyses using the HRS Psychosocial LBQ respondent weight yielded a meaningfully larger estimate: weighted HR, 1.17 (95% CI, 1.05-1.31; *P* =.005) compared with unweighted HR, 1.09 (95% CI, 0.99-1.20; *P* =.07) on the same subsample (*N* = 6808) (Table S11). This divergence may reflect differential selection into the LBQ sample or differences between conditional and marginal estimands. Because the lifelines robust sandwich estimator does not fully account for HRS primary sampling unit and stratum clustering, weighted standard errors may be anticonservative; accordingly, we retain the unweighted analysis as primary while reporting the weighted estimate as a sensitivity analysis.

Covariate-standardized absolute risks within age strata showed that among adults younger than 65 years, the adjusted 5-year mortality risk difference was 1.1 percentage points (5.5% vs 4.4%) and the 10-year difference was 2.3 percentage points (12.4% vs 10.1%). Among adults 65 years and older, adjusted risk differences were smaller (0.6 percentage points at 5 years, 1.0 percentage points at 10 years; Table S12). Accounting for household clustering (spouses) using robust standard errors yielded nearly identical point estimates with slightly wider confidence intervals (Table S13).

## Discussion

In this prospective cohort study of 7219 US adults aged 50 years and older with up to 16 years of follow-up, subjective financial strain was associated with a higher risk of incident self-reported heart disease (cause-specific HR, 1.18; 95% CI, 1.02-1.37; *P* =.03), an association that persisted after adjustment for household income and wealth and was corroborated by Fine-Gray subdistribution hazard models accounting for competing mortality (SHR, 1.16; 95% CI, 1.00-1.34; *P* =.04).

The mortality findings provide context: the association of financial strain with all-cause mortality was attenuated after adjustment for income and wealth, from HR, 1.17 (95% CI, 1.07-1.28) to HR, 1.10 (95% CI, 1.00-1.20; *P* =.051), a 41% reduction in excess risk. This attenuation was supported by propensity score matching (HR, 1.04; 95% CI, 0.91-1.19), modest E-values (CI-bound E-value, 1.00), and the absence of a monotonic dose-response gradient. The mortality association was significantly modified by age (*P* for interaction =.001), with HR, 1.25 (95% CI, 1.03-1.52) among adults younger than 65 years versus HR, 1.04 (95% CI, 0.94-1.16) among those 65 years or older. For clinical context, HRs in the 1.10 to 1.25 range are considerably smaller than those typically observed for established cardiometabolic risk factors in this age group (eg, current smoking, HR ∼2.0; diabetes, HR ∼1.5-1.8) and are closer in magnitude to other psychosocial exposures.

### Attenuation by Objective Socioeconomic Factors

The contrasting pattern between mortality and heart disease outcomes provides insight into the role of objective socioeconomic resources. For mortality, the association of financial strain largely overlapped with that captured by objective socioeconomic resources: adjustment for income and wealth reduced the HR from 1.17 (Model 3) to 1.10 (Model 3b), a 41% attenuation of excess risk. This finding is consistent with the strong SES-mortality gradient documented by Chetty et al^16^ but differs from some prior literature suggesting that subjective financial strain carries independent prognostic information beyond objective measures.^1^ For incident heart disease, however, the association with financial strain persisted after the same adjustment (HR, 1.18), suggesting that the cardiovascular association may not be fully accounted for by objective socioeconomic position.

Whether the mortality overlap reflects confounding or mediation is not identifiable from cross-sectional measurement of income, wealth, and financial strain. The attenuation from Model 3 to Model 3b should be interpreted as bounding the plausible range under different causal assumptions, not as evidence that objective resources causally “explain” the relationship. This interpretation is reinforced by the similarly attenuated results from propensity score matching (HR, 1.04; 95% CI, 0.91-1.19) and by the modest E-values indicating limited robustness to unmeasured confounding (CI-bound E-value, 1.00 for Model 3b; Table S3). Cross-classification by income quartile revealed a suggestive pattern of larger financial strain HRs at higher income levels (Q1, 0.99; Q4, 1.20), though all stratum-specific estimates had wide confidence intervals (Table S4).

### Time-Varying Exposure and Measurement Limitations

A first-order design limitation is the single-timepoint measurement of financial strain, which is inherently time-varying. The 2006 baseline assessment captures strain status at one moment in a dynamic economic environment; notably, the 2008 Great Recession occurred 2 years after baseline, likely altering the financial circumstances of many participants.

Nondifferential misclassification from this time-fixed measurement would be expected to attenuate hazard ratios toward the null, and the progressive attenuation with longer exclusion windows (HR, 1.08 excluding 1 year; HR, 1.05 excluding 2 years) is consistent with increasing exposure misclassification over time. Future studies could use repeated LBQ measurements across waves to model financial strain as a time-varying exposure.

### Age as an Effect Modifier

The association of financial strain with all-cause mortality was significantly modified by age (*P* for interaction =.001). The stronger association among adults younger than 65 years may reflect greater exposure to income volatility, caregiving responsibilities, and less comprehensive insurance before Medicare eligibility. At older ages, more stable income and health coverage may mitigate consequences of financial strain.

Alternative explanations include differential residual confounding by age, competing causes of death reducing the relative contribution of financial strain at older ages (diminishing marginal effects on relative hazards), healthy survivor bias attenuating associations among older adults, and differential exposure misclassification if financial strain is more volatile among working-age adults. The continuous age interaction confirmed a declining gradient from HR, 1.43 (95% CI, 1.21-1.70) at age 55 to HR, 0.95 (95% CI, 0.83-1.07) at age 80. Notably, no significant age interaction was observed for incident heart disease (Table 2), suggesting that the age-dependent pattern was specific to all-cause mortality and that noncardiovascular mechanisms may contribute to the mortality age interaction.

### Comparison With Prior Literature

Prior HRS research has linked objective financial measures to mortality and cardiovascular outcomes. Pool et al^8^ documented 50% higher mortality after acute wealth loss, Tucker-Seeley et al^9^ reported similar associations with material hardship, and Machado et al^10^ showed that downward wealth mobility during midlife was associated with worse long-term cardiovascular health. None of these studies examined whether subjective financial appraisal adds prognostic information beyond objective measures.

Szanton et al^17^ observed that perceived financial strain predicted mortality among community-dwelling older women even after adjusting for income, a result that contrasts with the substantial attenuation we observed; this discrepancy may reflect differences in population (women only, aged 65+), the financial strain measure (perceived income inadequacy versus ongoing stressor appraisal), or the adjustment strategy (income only versus income plus total net wealth). Samuel et al’s^18^ systematic review noted broadly consistent financial strain-mortality associations, but noted that few included studies adjusted for both income and wealth simultaneously. Berkowitz et al^19^ reported that financial strain was associated with increased health care expenditures independent of income, suggesting access-related pathways (such as deferred cardiovascular care or medication nonadherence) not fully captured by our adjustment.

### Strengths and Limitations

Strengths include the alignment between primary regression and propensity score models (allowing direct comparison of parametric and nonparametric estimates), use of the established HRS Psychosocial Leave-Behind chronic stressor measure, and sensitivity analyses including continuous age interaction testing, absolute risk estimation, and competing risks analysis for the cardiac outcome. The 16-year follow-up provides adequate power for rare outcomes, although it also introduces substantial exposure misclassification given the single-timepoint measurement of a time-varying exposure.

This study has several limitations. First, restricting to LBQ respondents with complete covariates may introduce selection bias; LBQ responders had modestly higher education than other Wave 8 respondents (SMD = 0.14; Table S9), and LBQ-weighted analyses yielded a larger HR (1.17 vs 1.09; Table S11), suggesting the unweighted primary estimate may be conservative relative to the population-level association. Weighted standard errors may be anticonservative, however, given HRS complex survey design limitations. Second, financial strain was measured at a single time point in 2006; subsequent changes, including the 2008 Great Recession, could produce substantial nondifferential misclassification, attenuating hazard ratios toward the null.

Third, residual confounding from unmeasured factors (eg, childhood socioeconomic conditions, negative affectivity, financial literacy) could explain the observed associations; the CI-bound E-value of 1.00 indicates that even weak unmeasured confounding could fully account for the primary mortality result. Fourth, reverse causation is possible; the attenuation with longer exclusion windows (HR, 1.08 at 1 year; HR, 1.05 at 2 years) may reflect reverse causation, progressive misclassification, or a time-limited early effect. Fifth, heart disease was self-reported using a composite measure with moderate validity (kappa approximately 0.47 vs Medicare claims), and detection bias is possible if financially strained individuals have different patterns of health care contact; as a secondary outcome, these measurement limitations warrant emphasis.

Sixth, the nonmonotonic dose-response pattern (“very upsetting” strain showing no mortality elevation; HR, 1.00; *n* = 373) raises construct validity questions, though this may reflect differential survivorship, safety-net engagement, or low statistical power in this small subgroup. Seventh, multiple comparisons were not formally adjusted; the cardiac *P* =.03 would not survive Bonferroni correction. Eighth, potential behavioral mediators (medication nonadherence, food insecurity, health literacy) were not assessed.

### Implications

These findings have implications for cardiovascular risk assessment. The association of financial strain with incident heart disease after adjustment for income and wealth supports consideration of financial strain screening in cardiovascular clinical practice. Asking about financial strain during cardiology visits or annual wellness visits could identify patients at elevated cardiovascular risk and prompt assessment of cost-related medication nonadherence, forgone preventive care, or food insecurity, all of which have established links to adverse cardiovascular outcomes.

For mortality, the attenuation after adjustment for objective socioeconomic resources suggests that financial strain functions primarily as a marker of socioeconomic disadvantage rather than an independent mortality risk factor. However, the age modification finding (*P* for interaction =.001), with HR, 1.25 (95% CI, 1.03-1.52) among adults younger than 65 years, suggests that financial strain screening may be most informative for preretirement adults who are not yet eligible for Medicare and are most likely to lack safety-net connections and stable health insurance coverage. Whether interventions addressing financial strain improve cardiovascular outcomes remains untested.

## Conclusions

In this cohort study, subjective financial strain was associated with incident heart disease independent of household income and wealth (cause-specific HR, 1.18; 95% CI, 1.02-1.37; *P* =.03). The association of financial strain with all-cause mortality was attenuated after adjustment for objective socioeconomic resources (HR, 1.10; 95% CI, 1.00-1.20; *P* =.051) and was modified by age (*P* for interaction =.001), with a significant association among adults younger than 65 years (HR, 1.25; 95% CI, 1.03-1.52) but not among those 65 years or older (HR, 1.04; 95% CI, 0.94-1.16). Absolute risk differences were small, and financial strain may serve as a clinically accessible marker of cardiovascular risk, particularly among preretirement adults.

Randomized trials of financial counseling, insurance navigation, or debt relief programs targeting cardiovascular outcomes are needed to determine whether addressing financial strain reduces incident heart disease among at-risk adults.

## Article Information

### Corresponding Author

Blinded for peer review.

### Author Contributions

Blinded for peer review.

## Acknowledgments

Generative AI tools were used to assist with code development, manuscript preparation, and editing. All analysis, interpretation, and conclusions are the sole responsibility of the author.

## Data Availability

The Health and Retirement Study data used in this analysis are publicly available from the Inter-university Consortium for Political and Social Research (ICPSR) at https://hrs.isr.umich.edu/. The RAND HRS Longitudinal File is available at https://www.rand.org/well-being/social-and-behavioral-policy/centers/aging/dataprod/hrs-data.html. Analysis code is available at https://github.com/DerekTharp/financial-strain-heart-disease.

## Supplemental Material

Tables S1-S13 Figures S1-S3 Data S1

## Nonstandard Abbreviations and Acronyms

Abbreviation: Definition
ATT: average treatment effect on the treated
CES-D: Center for Epidemiologic Studies Depression Scale
EFTF: enhanced face-to-face interview
HRS: Health and Retirement Study
LBQ: Leave-Behind Questionnaire
SHR: subdistribution hazard ratio
SMD: standardized mean difference

---

**Figure 1.**
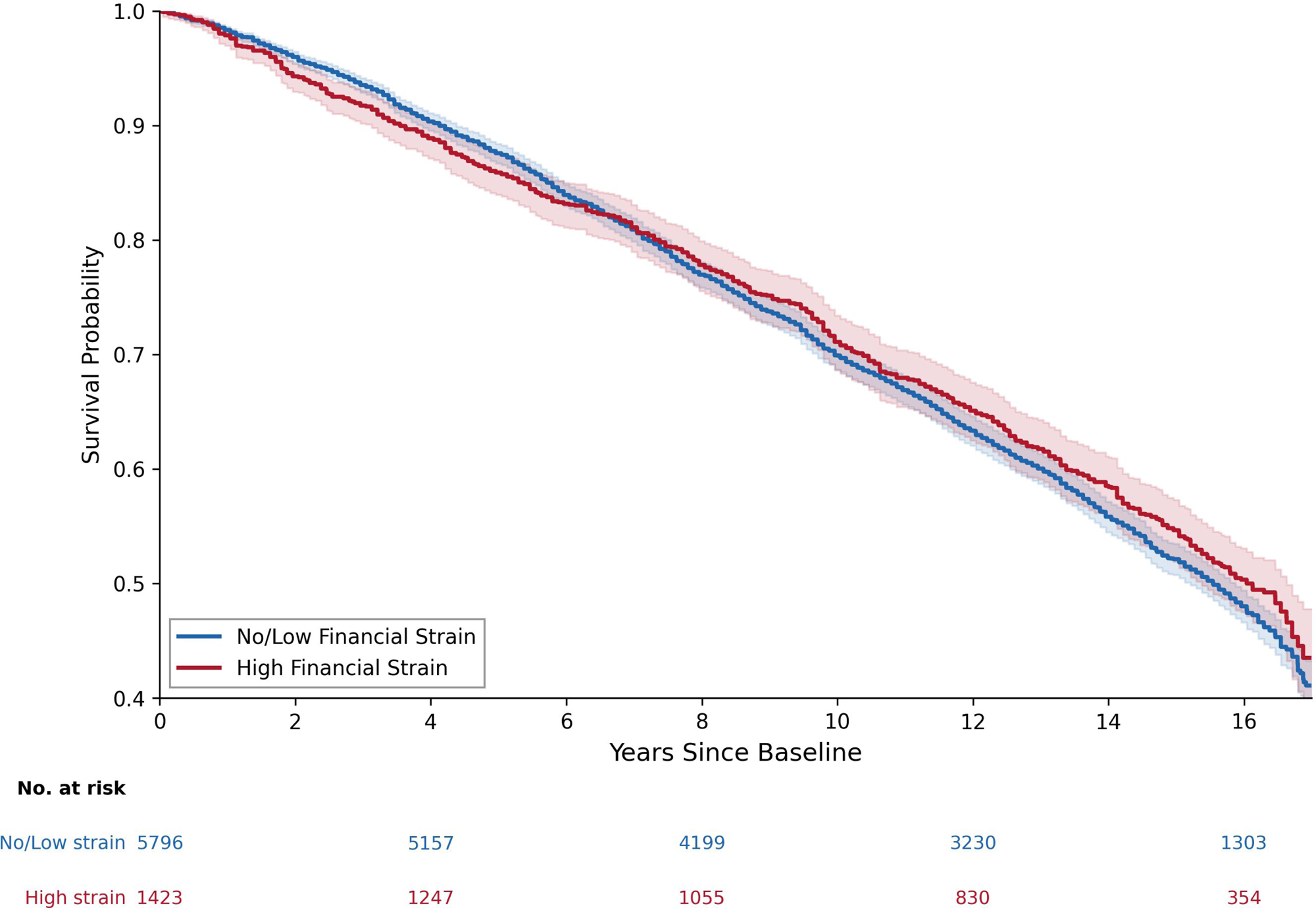
Kaplan-Meier Survival Curves for All-Cause Mortality by Financial Strain Status. Survival probability over 16 years of follow-up among 7219 adults aged 50 years and older, stratified by financial strain status. Shaded areas indicate 95% CIs. Log-rank test *P* =.20.

**Figure 2.**
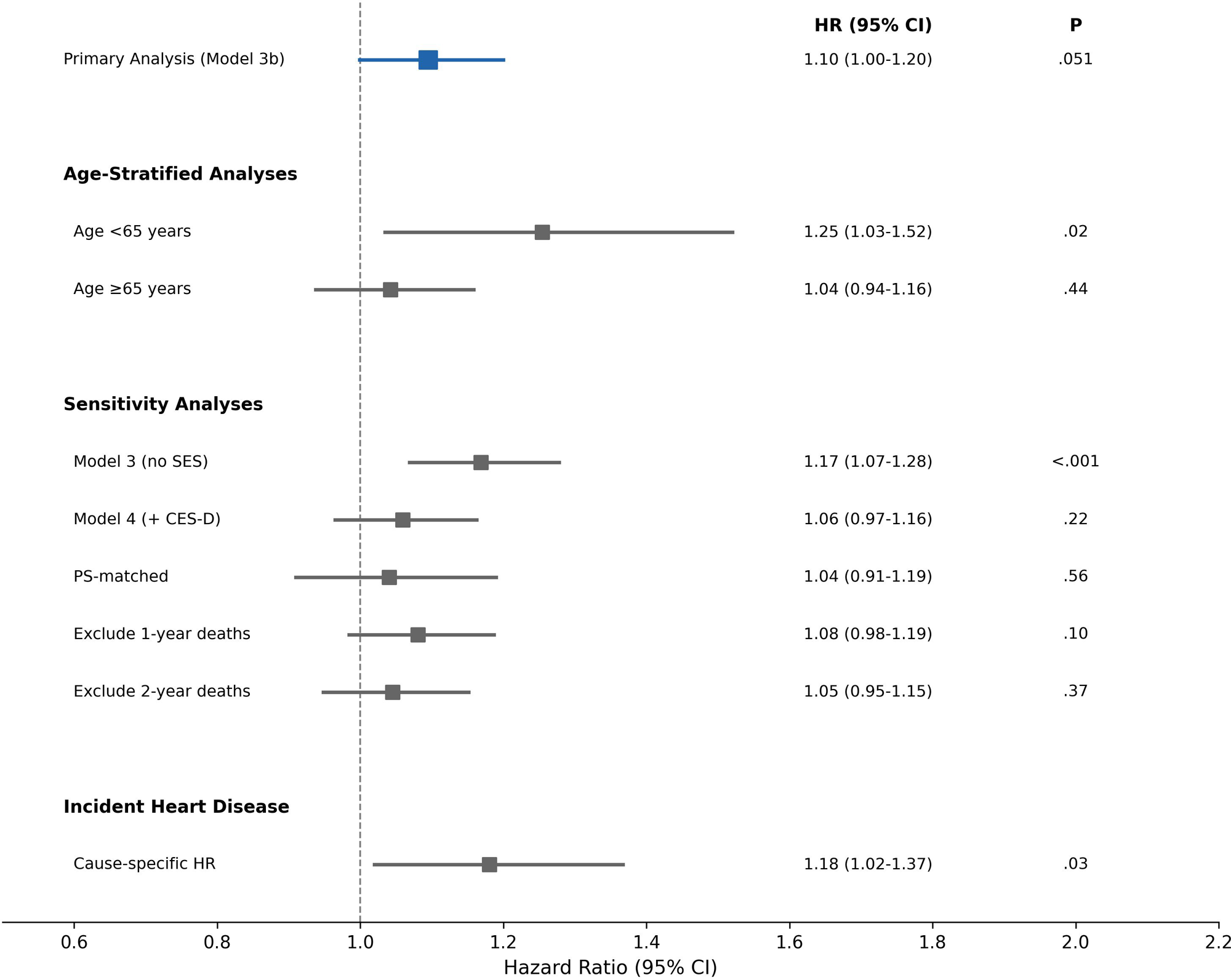
Forest Plot of Financial Strain-Mortality Associations Hazard ratios and 95% CIs for the association of financial strain with all-cause mortality across primary and sensitivity analyses. The primary analysis (Model 3b) is highlighted; sensitivity analyses are shown for comparison.

## References

1. Kahn JR, Pearlin LI. Financial strain over the life course and health among older adults. J Health Soc Behav. 2006;47:17–31.

2. Sinclair RR, Cheung JH. Money matters: recommendations for financial stress research in occupational health psychology. Stress Health. 2016;32:181–193.

3. Zimmerman FJ, Katon W. Socioeconomic status, depression disparities, and financial strain: what lies behind the income-depression relationship? Health Econ. 2005;14:1197–1215.

4. McEwen BS. Stress, adaptation, and disease: allostasis and allostatic load. Ann N Y Acad Sci. 1998;840:33–44.

5. Steptoe A, Kivimaki M. Stress and cardiovascular disease. Nat Rev Cardiol. 2012;9:360–370.

6. Epel ES, Blackburn EH, Lin J, et al. Accelerated telomere shortening in response to life stress. Proc Natl Acad Sci U S A. 2004;101:17312–17315.

7. Francoeur RB. Cumulative financial stress and strain in palliative radiation outpatients: the role of age and disability. Acta Oncol. 2005;44:369–381.

8. Pool LR, Burgard SA, Needham BL, Elliott MR, Langa KM, Mendes de Leon CF. Association of a negative wealth shock with all-cause mortality in middle-aged and older adults in the United States. JAMA. 2018;319:1341–1350.

9. Tucker-Seeley RD, Li Y, Subramanian SV, Sorensen G. Financial hardship and mortality among older adults using the 1996-2004 Health and Retirement Study. Ann Epidemiol. 2009;19:850–857.

10. Machado S, Gallo WT, Engel L. Midlife wealth mobility and long-term cardiovascular health. J Am Heart Assoc. 2021;10:e019293.

11. Sonnega A, Faul JD, Ofstedal MB, Langa KM, Phillips JW, Weir DR. Cohort profile: the Health and Retirement Study (HRS). Int J Epidemiol. 2014;43:576–585.

12. Health and Retirement Study. 2006 Psychosocial Leave-Behind Questionnaire. Survey Research Center, Institute for Social Research, University of Michigan. Accessed February 2, 2025. https://hrs.isr.umich.edu

13. Austin PC. An introduction to propensity score methods for reducing the effects of confounding in observational studies. Multivariate Behav Res. 2011;46:399–424.

14. Rosenbaum PR, Rubin DB. The central role of the propensity score in observational studies for causal effects. Biometrika. 1983;70:41–55.

15. VanderWeele TJ, Ding P. Sensitivity analysis in observational research: introducing the E-value. Ann Intern Med. 2017;167:268–274.

16. Chetty R, Stepner M, Abraham S, et al. The association between income and life expectancy in the United States, 2001-2014. JAMA. 2016;315:1750–1766.

17. Szanton SL, Allen JK, Thorpe RJ Jr, Seeman T, Bandeen-Roche K, Fried LP. Effect of financial strain on mortality in community-dwelling older women. J Gerontol B Psychol Sci Soc Sci. 2008;63:S369–S374.

18. Samuel LJ, Commodore-Mensah Y, Engel L, Gallo JJ, Szanton SL. Subjective financial strain and mortality: a systematic review. J Epidemiol Community Health. 2025;79:187–195.

19. Berkowitz SA, Seligman HK, Meigs JB, Basu S. Food insecurity, health care utilization, and high cost: a longitudinal cohort study. Am J Manag Care. 2018;24:399–404.

